# Association of COVID-19 incidence with objectively and subjectively measured mental health proxies in the Austrian Football League – an epidemiological study

**DOI:** 10.1101/2021.01.27.21250527

**Authors:** Antje van der Zee-Neuen, Alexander Seymer, Dagmar Schaffler-Schaden, Jürgen Herfert, James O’Brien, Tim Johansson, Patrick Kutschar, Stephan Ludwig, Thomas Stöggl, David Keeley, Maria Flamm, Jürgen Osterbrink

## Abstract

**Objective:** We aimed to explore the association of COVID-19 incidence with mental health in 225 team and staff members of five professional Austrian Football clubs captured by objective (location variance) or subjective (self-reported sleep quality, level of recovery, perceived risk of infection) mental health proxies.

**Methods:** Data collected during the implementation of a novel monitoring concept to enable safe continuation of professional Football during the COVID-19 pandemic were matched with Austrian COVID-19 incidence data and smartphone collected location data (time-period June 17^th^ to July 31^st^, 2020). Multivariable linear regression models explored the association of COVID-19 incidence, defined as daily novel or active cases of COVID-19, with the objective and subjective health proxies while adjusting for the occurrence of one COVID-19 case in a staff member in one of the clubs, team status (i.e. player vs staff) and game days.

**Results:** Data from 115 participants were analysed. An increasing number of novel COVID-19 cases was significantly associated with deteriorating sleep quality (B 0.48, 95% CI 0.05; 1.00) but with none of the other mental health proxies. An increasing number of active COVID-19 cases was significantly associated with an increase in perceived infection risk (B 0.04, 95% CI 0.00; 0.07) and location variance (B 0.28, 95% CI 0.06; 0.49).

**Conclusion:** The adverse association of an increasing COVID-19 incidence with mental health in professional Footballers and staff members became obvious particularly in subjectively measured mental health. During the ongoing pandemic, targeted mental care should be included in the daily routines of this population.

**SUMMARY BOX**

- An increasing COVID-19 incidence is associated with deterioration of mental health in players and staff members of professional Football teams.
- The perceived COVID-19 infection risk is more pronounced in staff members than in players of professional Football clubs.
- Surprisingly, a rise in the number of active COVID-19 cases results in an increasing location variance.

## INTRODUCTION

The COVID-19 pandemic is affecting not only physical health and societal structures, but also the mental health of individuals facing the burden of necessary alterations to daily life (1, 2). Quarantine and related isolation have influenced the mental wellbeing of individuals confronted with previous epidemics (3–5). The duration of the current pandemic and preventive measures for disease-transmission (like social distancing, isolation and quarantine) might result in an additional rise of the substantial burden of mental health disorders (1, 6). Moreover, daily news reports on infection and mortality rates may be perceived as threatening leading to anxiety and stress. Therefore, measures to enhance the early detection of mental health problems are essential. The monitoring of physical activity (PA) offers a subtle way for this purpose as reduced PA predicts the occurrence of depressive symptoms (7, 8). Self-reported PA, activity trackers or interviewing by caretakers in the general population or in populations under medical supervision can aid in the subtle detection of mental health issues (9). This appears less useful for professional athletes, as levels of PA are influenced by training routines, competitions and recovery periods. Professional athlete’s (mental) health is generally well monitored by their accompanying (para-) medical staff. However, stigmatization and reluctance to discuss mental health problems might inhibit early detection of the latter (10, 11). Yet, professional athletes might be at increased risk of developing such problems due to considerable stress in relation to both, their professional activities <u>and</u> the COVID-19 pandemic. This emphasizes the necessity of exploring possibilities for the monitoring of athlete’s mental health, preferably unobtrusively. Emerging evidence suggests that individual mobility measured by smartphones may successfully detect adverse alterations in mental health (12, 13).

The resumption of professional sports during the COVID-19 pandemic required the close monitoring of athletes’ health to prevent disease transmission among team and staff members. In Austria, the monitoring of five professional Football clubs included smartphone-based geo-tracking, allowing for the estimation of risk exposure of footballers outside the training and competition facilities (14). However, the geo-tracking data also yielded the opportunity of exploring potential mental health problems in participants. A previous study by Saeb et al. found that less location variance (obtained by means of smartphone mobility tracing) correlated with deteriorating mental health, specifically with depression (12).

In the current study, we aimed to explore the association of COVID-19 incidence with mental health in team and staff members of five professional Austrian football clubs captured by objective (i.e. location variance) or subjective (i.e. self-reported sleep quality, level of recovery and perceived risk of infection) proxies. We hypothesized, that adverse alterations in mental health would be evident for the objective proxy but not for the subjective proxies (given the reluctance to reveal mental health problems) and, that an increasing COVID-19 incidence would result in lower levels of location variance.

## METHODS

In the context of the scientific surveillance of the mandatory monitoring concept of the Austrian Football Association across four Premier League clubs and one Second League club (i.e. club A-club E), collection of geo-tracking and health diary data took place during a 12-week observation period. Players and staff members were included. The current study presents secondary analyses of these data (14).

### Patient or Public Involvement

Patients or the public were not involved in the design, conduct, reporting or dissemination plans of our research.

### Smartphone – App

The smartphone application (app) “Wallpass™” (developed by the US-based company Electronic Caregiver) collected geo-tracking data. After installation on the primary mobile device of participants, the app collected motion triggered Global Navigation Satellite System (GNSS) data points. In a second step, machine-learning algorithms assigned activities and locations to the GNSS data. The data was stored in flexible schemes of a noSQL database, in order to gather the multiple layers of information associated with each data point. Participants received unique identification-codes for the installation of the app. Club physiotherapists kept a list of codes and corresponding names. Researchers did not have access to the names corresponding with the identification-codes.

### Parameters

In the current study, several parameters were included that were part of the health diaries of the original study, calculated based on the collected geo-tracking data or provided by the Austrian government.

### Self-reported mental health proxies (i.e. subjective proxies)

#### Sleep quality

As impairments in sleep quality are linked to decreased mental health (15), this parameter was included as mental health proxy (10-point scale; 1=good sleep, quickly fell asleep slept through the night, 10=bad sleep, problems falling asleep, several night time wake-ups).

### Level of recovery

An athlete’s recovery level is associated with alterations in mood (16) and was therefore installed as proxy indicator for mental health (10-point scale; 1=energetic, cheerful, rested; 10=tired, unenthusiastic, exhausted).

### Subjective risk of infection

The current perceived risk of infection through COVID-19 was used as indicator for anxiety related to the pandemic (10-point scale; 1=calm, composed, unconcerned; 10=panicked, anxious, severe risk).

#### Mental health proxy measured through smartphone mobility tracing (i.e. objective proxies)

### Location variance

Location variance describes participant mobility in terms of GNSS-location variance independent of type of location (i.e. variability in GNSS location). In the current study, this parameter was included as objective mental health proxy and calculated as suggested by Saeb et al (12). The first step in the calculation involved reversing geocoding (i.e. assignment of ZIP-codes to raw GNSS data). For this purpose, the free online services Photon API and BING API were used (17, 18). Next, location variance was calculated while accounting for distribution skewness in location data across subjects by means of the logarithm of the sum of the statistical variances of latitude and longitude components of the daily location data.

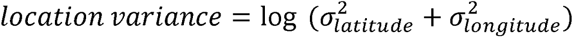

#### COVID-19 incidence data

The Austrian National Public Health Institute (Gesundheit Österreich GmbH, GÖG) is continuously monitoring the occurrence of COVID-19 cases (19). For the current investigation, they provided access to the anonymized database of the electronic epidemiological reporting system (EMS). The database offered case-specific information on the date of diagnosis, (potential) date of death, (potential) date of recovery from COVID-19 and geographical region of occurrence in terms of 33 local administrative districts listed in supplementary table S1. The daily number of COVID-19 cases was determined by means of date aggregation. Two parameters were then computed based on the aggregated data: 1) number of <u>**novel**</u> COVID-19 cases per day and 2) <u>**total**</u> number of <u>**active**</u> COVID-19 infections per day.

### Additional parameters for adjustment

#### COVID-19 case in one Football-club

In one of the participating Football clubs, a COVID-19 case occurred during the observation period. The COVID-19 case was not part of the monitored sample, but an employee working in club administration. Due to inter-club co-operation, this individual had contact with two of the five teams in the present study.

As the occurrence of a case in proximity to participants might affect the association of mental health and COVID-19 incidence, we constructed a variable for adjustment comprising three categories:

1. all clubs without known COVID-19 incidence in their proximity (not in proximity of case)
2. clubs in proximity of COVID-19 case before occurrence (clubs before COVID-19 case)
3. clubs in in proximity of COVID-19 case after occurrence (clubs after COVID-19 case)

#### Team status

Individuals participating in the study were either players or staff members of the participating clubs. To account for potential differences in the mental health proxies attributable to being part of any of these two groups, the binary variable team status was computed.

#### Game day

Existing evidence suggests that competitions in professional athletes are related to increased levels of anxiety and stress (20). In addition, game days inevitably participants’ movement due to travel to and from competition. Therefore, a variable differentiating between game and no-game days during the observation period was computed.

### Statistical analyses

All analyses were performed using R 4.0.3 (21). Analyses were restricted to complete data only. A value of p≤0.05 was considered statistically significant.

#### Data

The players and staff of five clubs reported 120,938 GNSS data points via the smartphone app from June 17^th^ to July 31^st^, 2020. Theoretically, 225 players and staff members were supposed to participate but only 115 unique user ids were recorded by the app. The data provided by each of the 115 users differed significantly leading to a highly skewed distribution of GNSS data points per user (Figure 1). Supplementary figure S1 provides a more detailed illustration of the distribution.

**Figure 1.**
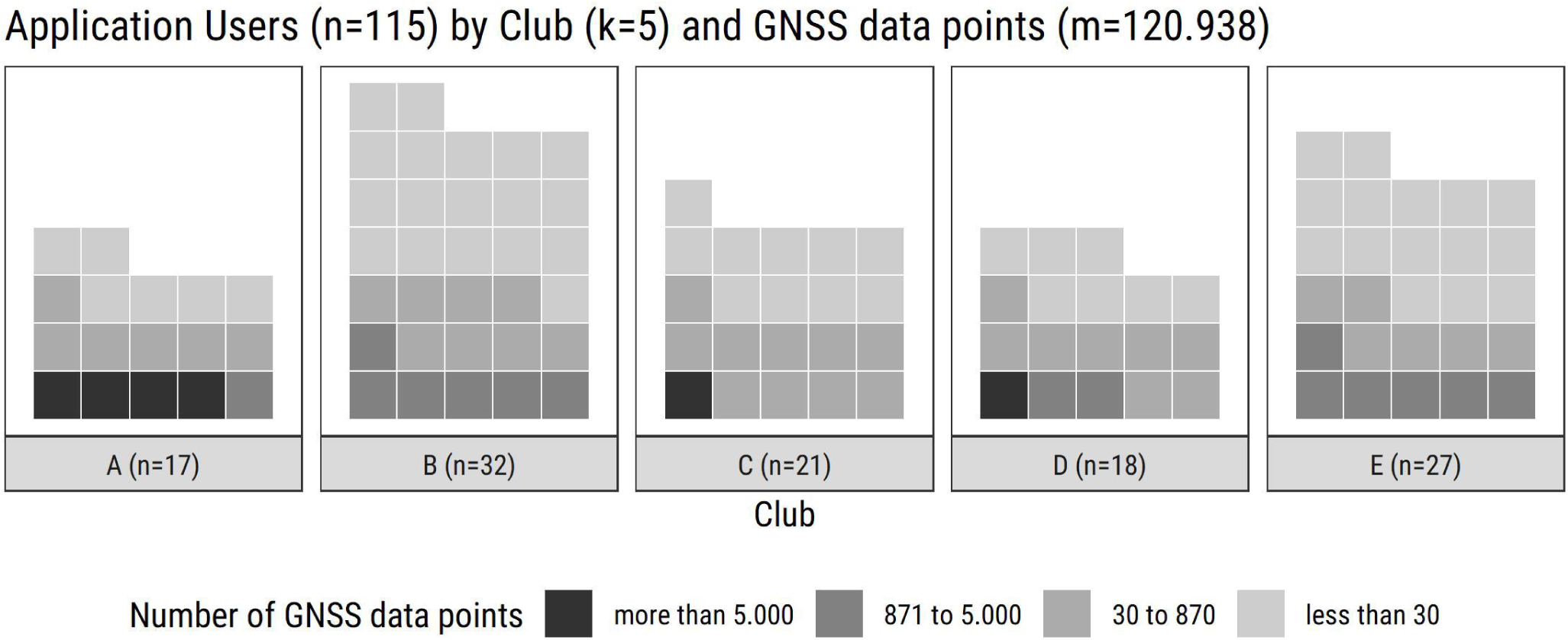
Distribution of Global Navigation Satellite System (GNSS) data points in participating clubs *Note: As our analysis required data aggregation, a minimum of 30 GNSS data points a day would be the bare minimum to be considered in the later analysis. And with 29 days between first data collection (June 17*^*th*^*) and final competition in the premier leagues season (July 15*^*th*^*) a full dataset with 30 GNSS data points a day would require 870 GNSS data points in total*.

As COVID-19 incidence data was provided only on the local administrative districts, GNSS location data were matched with these care regions based on the previously identified postcodes. Next, self-reported mental health proxy indicators were included in the same data set based on participant-specific identification-codes and reported day. The 120,938 GNSS data points were aggregated by unique user id and date resulting in a user-day-dataset with 508 cases (Supplementary figure S1). Cases aggregated with less than 30 GNSS data points were excluded leaving 297 user-day-data points left for the analysis. All results rest on these 297 user-day data points from 57 app users.

#### Descriptive statistics

All variables included in the current study were described for user-day cases according to their metric properties (i.e. means, standard deviation (SD), median, minimum and maximum for continuous data and frequencies (n) and percentages (%) for categorical data). A single user-day case is the unique combination of user and day during the observation period (e.g. a single user day-case for new COVID-19 cases is the mean of the official number of new cases in the region visited by the included users on a specific day weighted for the number of data points per visited region).

#### Association of incidence of COVID-19 with mental health proxies

First, bivariate correlations (Pearson’s r) explored the association of novel and total COVID-19 incidence with the objective and subjective mental health proxies.

Subsequently, multivariable linear regression models with ordinary least square estimation explored the association of the COVID-19 incidence variables with the objective (location variance) and subjective mental health proxies (sleep quality, level of recovery, perceived risk of infection), while adjusting for game day, team status and the occurrence of a COVID-19 case in one participating club.

## RESULTS

The sample for further analyses consisted of 297 cases. The sample consisted of 205 cases from staff members (69%) and 92 cases from players (31%). Table 1 provides an overview of user-day cases for the total sample and each participating club.

**Table 1.** Characteristics of user-day cases (n) for the total sample and each participating club

|  | *Club A*<br>*(n=66)* | *Club B*<br>*(n=93)* | *Club C*<br>*(n=42)* | *Club D*<br>*(n=32)* | *Club E*<br>*(n=64)* | *Overall*<br>*(n=297)* |
| --- | --- | --- | --- | --- | --- | --- |
| *Sleep quality* |  |  |  |  |  |  |
| Mean (SD) | 2.1 (1.2) | 2.92 (1.6) | 2.5(2.3) | NA (NA) | 1.8 (0.8) | 2.3 (1.6) |
| Median [Min, Max] | 2.0 [1.0, 5.0] | 2.0 [1.0, 7.0] | 2.0 [1.0, 10.0] | NA [NA, NA] | 2.0 [1.0, 3.0] | 2.0 [1.0, 10.0] |
| Missing | 36 (54.5%) | 69 (74.2%) | 13 (31.0%) | 32 (100%) | 35 (54.7%) | 185 (62.3%) |
| *Recovery* |  |  |  |  |  |  |
| Mean (SD) | 2.0 (1.1) | 2.9 (1.4) | 2.9 (2.1) | NA (NA) | 2.5 (1.1) | 2.5 (1.5) |
| Median [Min, Max] | 2.0 [1.0, 5.0] | 3.0 [1.0, 6.0] | 2.0 [1.0, 8.0] | NA [NA, NA] | 3.0 [1.0, 5.0] | 2.0 [1.0, 8.0] |
| Missing | 36 (54.5%) | 72 (77.4%) | 13 (31.0%) | 32 (100%) | 35 (54.7%) | 188 (63.3%) |
| *Perceived risk of infection* |  |  |  |  |  |  |
| Mean (SD) | 1.5 (0.8) | 1.4 (0.7) | 1.2 (0.5) | NA (NA) | 1.0 (0.0) | 1.3 (0.6) |
| Median [Min, Max] | 1.0 [1.0, 3.0] | 1.0 [1.0, 3.0] | 1.0 [1.0, 3.0] | NA [NA, NA] | 1.0 [1.0, 1.0] | 1.0 [1.0, 3.0] |
| Missing | 36 (54.5%) | 80 (86.0%) | 13 (31.0%) | 32 (100%) | 35 (54.7%) | 196 (66.0%) |
| *Location Variance\** |  |  |  |  |  |  |
| Mean (SD) | -16.6 (9.2) | -24.4 (14.7) | -14.9 (7.5) | -17.5 (10.9) | -17.7 (11.7) | -19.1 (12.2) |
| Median [Min, Max] | -15.5 [-52.6, -2.2] | -19.3 [-56.0, -2.8] | -15.5 [-48.4, 0.0] | -15.2 [-46.7, -1.8] | -14.2 [-47.3, -1.1] | -16.0 [-56.0, 0.0] |
| *Novel COVID-19 cases per day* |  |  |  |  |  |  |
| Mean (SD) | 0.8 (0.6) | 0.9 (0.6) | 1.3 (0.9) | 0.3 (0.3) | 0.8 (0.7) | 0.9 (0.7) |
| Median [Min, Max] | 0.7 [0.0, 2.5] | 0.828 [0, 3.67] | 1.26 [0, 3.60] | 0.3 [0.0, 1.4] | 0.7 [0.0, 2.7] | 0.8 [0.0, 3.7] |
| Missing | 3 (4.5%) | 0 (0%) | 2 (4.8%) | 3 (9.4%) | 26 (40.6%) | 34 (11.4%) |
| *Total active COVID-19 cases per day* |  |  |  |  |  |  |
| Mean (SD) | 11.1 (6.0) | 12.8 (7.0) | 16.6 (6.2) | 2.9 (2.3) | 7.4 (4.1) | 11.1 (7.02) |
| Median [Min, Max] | 8.7 [4.3, 34.5] | 14.9 [2.8, 31.2] | 17.7 [0.0, 24.8] | 1.8 [1.8, 10.0] | 6.5 [0.9, 18.7] | 9.2 [0.0, 34.5] |
| Missing | 3 (4.5%) | 0 (0%) | 2 (4.8%) | 3 (9.4%) | 26 (40.6%) | 34 (11.4%) |
| *COVID-19 occurrence<sup>1</sup>, n (%)* |  |  |  |  |  |  |
| Clubs before COVID-19 case | 0 (0.0%) | 0 (0.0%) | 0 (0.0%) | 0 (0.0%) | 13 (20.3%) | 13 (4.4%) |
| Clubs after COVID-19 case | 66 (100.0%) | 0 (0.0%) | 0 (0.0%) | 0 (0.0%) | 51 (79.7%) | 117 (39.4%) |
| not in proximity of case | 0 (0.0%) | 93 (100.0%) | 42 (100.0%) | 32 (100.0%) | 0 (0.0%) | 167 (56.2%) |
| *Team status, n(%)* |  |  |  |  |  |  |
| Staff | 37 (56.1%) | 80 (86.0%) | 9 (21.4%) | 28 (87.5%) | 51 (79.7%) | 205 (69.0%) |
| Player | 29 (43.9%) | 13 (14.0%) | 33 (78.6%) | 4 (12.5%) | 13 (20.3%) | 92 (31.0%) |
| *Game day, n(%)* |  |  |  |  |  |  |
| Yes | 10 (15.2%) | 13 (14.0%) | 7 (16.7%) | 6 (18.8%) | 9 (14.1%) | 45 (15.2%) |
| No | 56 (84.8%) | 80 (86.0%) | 35 (83.3%) | 26 (81.2%) | 55 (85.9%) | 252 (84.8%) |

\* Lower values represent larger variance in location
<sup>1</sup> In one of the participating Football-clubs, a COVID-19 infection occurred during the observation period. The categories of this variable distinguishes the cases before and after this infection and cases that were not in the proximity of cases.

### Association of incidence of COVID-19 with mental health parameters

Location variance correlated with both COVID-19 case variables. Furthermore, novel COVID-19 cases per day were significantly associated with sleep quality, while the total number of active COVID-19 cases showed a relationship with the level of recovery instead. All three subjective indicators related to each other (Table 2). Supplementary figure S2 provides a more detailed illustration of correlation.

**Table 2.** Correlation matrix of metric parameters

|  | Sleep quality | Level of recovery | Perceived infection risk | Location Variance | Novel COVID-19 cases per day | Total active COVID-19 cases per day |
| --- | --- | --- | --- | --- | --- | --- |
| Sleep quality |  | 0.555<sup>*</sup> | 0.313<sup>*</sup> | 0.151 | 0.242<sup>*</sup> | 0.194 |
| Level of recovery |  |  | 0.309<sup>*</sup> | 0.061 | 0.193 | 0.244<sup>*</sup> |
| Perceived risk of infection |  |  |  | 0.120 | 0.052 | 0.090 |
| Location Variance |  |  |  |  | 0.128<sup>*</sup> | 0.177<sup>*</sup> |
| Novel COVID-19 cases per day |  |  |  |  |  | 0.637<sup>*</sup> |
| Total active COVID-19 cases per day |  |  |  |  |  |  |
| Computed correlation used Pearson-method with pairwise-deletion; * p≤0.05 |  |  |  |  |  |  |

Multivariable linear regression models exploring the association of novel COVID-19 infections with mental health proxies showed a significant association of novel COVID-19 cases per day with deterioration of sleep quality (B 0.48, 95% CI 0.05; 1.00) but with none of the other subjective and objective mental health proxies. When compared to staff members, players had a significantly lower perceived risk of infection (B −0.26, 95% CI −0.52; −0.00) and a significantly higher variance in their location (B 5.17, 95%CI 2.04; 8.29). Moreover, the perceived risk of infection was significantly higher after a COVID-19 case occurred in one of the clubs (B 0.52, 95% CI 0.06; 0.98). The model including the perceived risk of infection as outcome variable showed the best fit (adjusted R^2^=0.079, F=2.440, df=5, p=0.041). (Table 3)

**Table 3.**
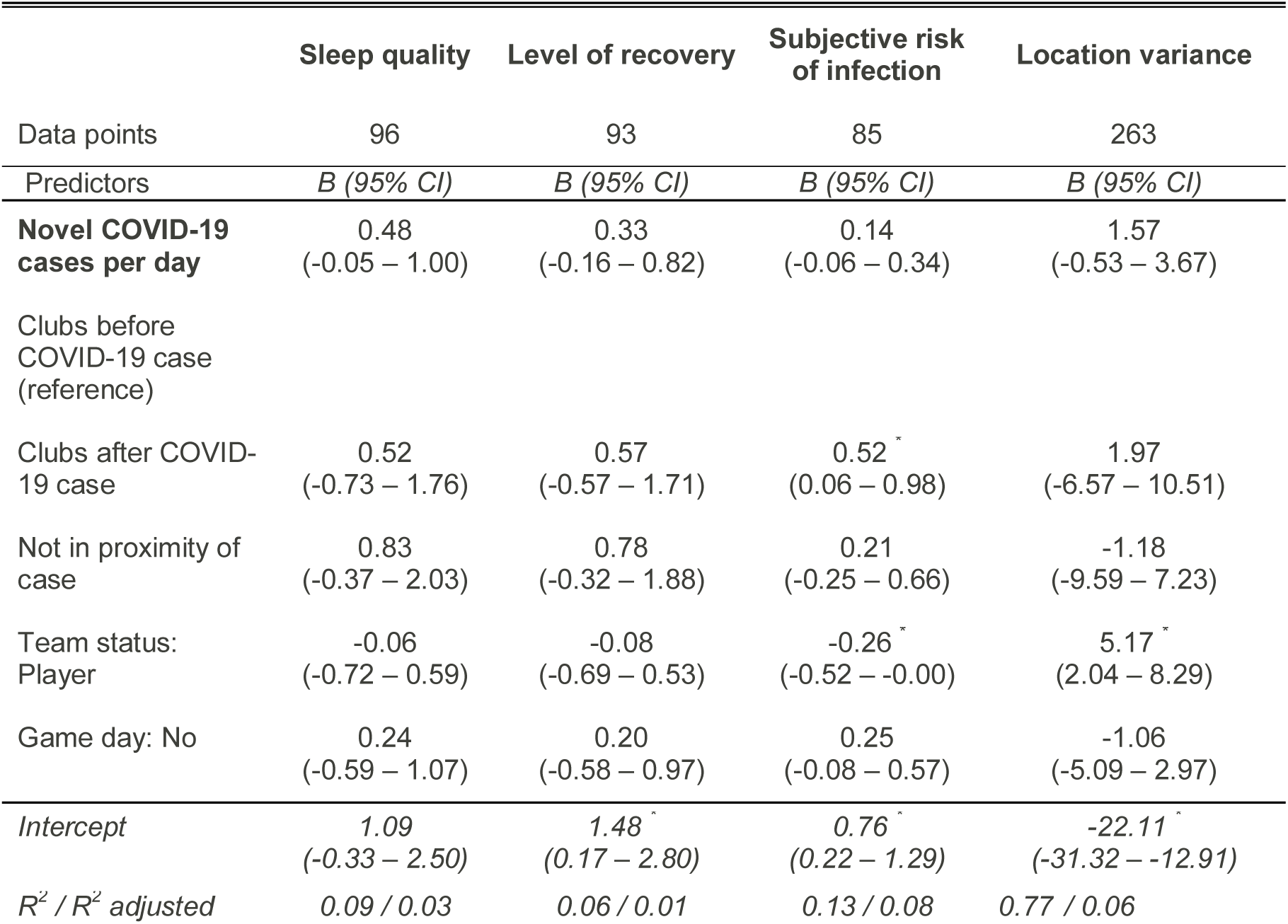
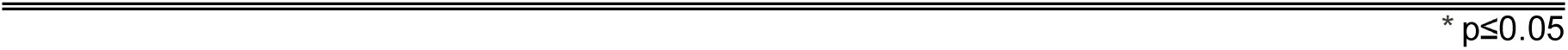
Multivariable linear regression models exploring the association of <u>novel COVID-19 cases</u> per day with mental health proxies

|  | Sleep quality | Level of recovery | Subjective risk of infection | Location variance |
| --- | --- | --- | --- | --- |
| Data points | 96 | 93 | 85 | 263 |
| Predictors | *B* (95% *CI*) | *B* (95% *CI*) | *B* (95% *CI*) | *B* (95% *CI*) |
| **Novel COVID-19 cases per day** | 0.48<br>(-0.05 – 1.00) | 0.33<br>(-0.16 – 0.82) | 0.14<br>(-0.06 – 0.34) | 1.57<br>(-0.53 – 3.67) |
| Clubs before COVID-19 case (reference) |  |  |  |  |
| Clubs after COVID-19 case | 0.52<br>(-0.73 – 1.76) | 0.57<br>(-0.57 – 1.71) | 0.52<sup>*</sup><br>(0.06 – 0.98) | 1.97<br>(-6.57 – 10.51) |
| Not in proximity of case | 0.83<br>(-0.37 – 2.03) | 0.78<br>(-0.32 – 1.88) | 0.21<br>(-0.25 – 0.66) | -1.18<br>(-9.59 – 7.23) |
| Team status: Player | -0.06<br>(-0.72 – 0.59) | -0.08<br>(-0.69 – 0.53) | -0.26<sup>*</sup><br>(-0.52 – -0.00) | 5.17<sup>*</sup><br>(2.04 – 8.29) |
| Game day: No | 0.24<br>(-0.59 – 1.07) | 0.20<br>(-0.58 – 0.97) | 0.25<br>(-0.08 – 0.57) | -1.06<br>(-5.09 – 2.97) |
| *Intercept* | 1.09<br>(-0.33 – 2.50) | 1.48<sup>*</sup><br>(0.17 – 2.80) | 0.76<sup>*</sup><br>(0.22 – 1.29) | -22.11<sup>*</sup><br>(-31.32 – -12.91) |
| *R*<sup>2</sup> / *R*<sup>2</sup> adjusted | 0.09 / 0.03 | 0.06 / 0.01 | 0.13 / 0.08 | 0.77 / 0.06 |

In the models exploring the association of the total number of active COVID-19 infections with the mental health proxies, an increasing number of active COVID-19 cases was significantly associated with an increased perceived risk of infection (B 0.04, 95% CI 0.00; 0.07) and surprisingly with an increase in the location variance (B 0.28, 95% CI 0.06; 0.49). As in the models including novel COVID-19 infections as main independent variable of interest, players had a significantly lower perceived risk of infection when compared to staff members and a significantly higher variance in their location. Moreover, the model including the perceived risk of infection as outcome variable showed the best fit (adjusted R^2^=0.112, F=3.126, df=5, p=0.013). (Table 4)

**Table 4.** Multivariable linear regression models exploring the association of total active COVID-19 cases per day with mental health proxies

|  | Sleep quality | Level of recovery | Subjective risk of infection | Location variance |
| --- | --- | --- | --- | --- |
| Data points | 96 | 93 | 85 | 263 |
| Predictors | B (95%CI) | B (95%CI) | B (95%CI) | B (95%CI) |
| **Total active COVID-19 cases per day** | 0.02<br>(-0.05 – 0.09) | 0.05<br>(-0.02 – 0.12) | 0.04 *<br>(0.00 – 0.07) | 0.28 *<br>(0.06 – 0.49) |
| Clubs before COVID-19 case (reference) |  |  |  |  |
| Clubs after COVID-19 case | 0.40<br>(-0.93 – 1.73) | 0.29<br>(-0.91 – 1.49) | 0.34<br>(-0.14 – 0.82) | 0.12<br>(-8.49 – 8.73) |
| Not in proximity of case | 0.77<br>(-0.74 – 2.28) | 0.28<br>(-1.11 – 1.68) | -0.19<br>(-0.78 – 0.41) | -3.39<br>(-11.96 – 5.18) |
| Team status: Player | 0.02<br>(-0.65 – 0.68) | -0.05<br>(-0.66 – 0.55) | -0.27 *<br>(-0.52 – -0.01) | 4.75 *<br>(1.65 – 7.85) |
| Competition: No | 0.27<br>(-0.57 – 1.12) | 0.21<br>(-0.57 – 0.99) | 0.27<br>(-0.05 – 0.59) | -1.24<br>(-5.24 – 2.76) |
| (Intercept) | 1.28<br>(-0.14 – 2.71) | 1.53 *<br>(0.23 – 2.83) | 0.72 *<br>(0.20 – 1.25) | -21.54 *<br>(-30.63 – -12.45) |
| R2 / R2 adjusted | 0.06 / 0.00 | 0.07 / 0.01 | 0.17 / 0.11 | 0.92 / 0.07 |

\* p≤0.05

## DISCUSSION

To our knowledge, this is the first study exploring the association of COVID-19 incidence with subjective and objective proxies for mental health in elite Footballers and their professional surrounding. Contrary to our hypotheses, adverse alterations in mental health became particularly evident for the subjective mental health proxies. Moreover, we found that an increasing number of novel cases of COVID-19 was significantly associated with worse sleep quality and that an increasing total of active COVID-19 cases was significantly associated with higher perceived risk of infection and with a higher variance in location. The latter is particularly surprising, as we assumed that, with increasing COVID-19 incidence, the location variance would decrease due to risk avoidance behaviour (22). This finding might be attributable to the low incidence of COVID-19 during the observation period (23, 24) but may also suggest that location variance is not suited to capture mental health alterations due to COVID-19. In this line, the perceived risk of infection did in fact increase but no significant correlation of this variable with location variance was found, indicating the independence of these constructs.

The adverse association of COVID-19 with sleep quality is in line with previous research findings focusing on the general population and professional athletes (25–27). In an international study across 49 countries 40% of participants reported worse sleep quality than before the pandemic and the consumption of sleeping pills increased by 20% when compared to the pre-pandemic situation. According to the study, the deterioration in sleep was among others attributable to quarantine measures and the adverse impact of the pandemic on participant’s livelihood (28). In the current study, the perceived risk of infection was included as a proxy for anxiety. Sjöberg defined perceived risk as an individuals’ psychological evaluation of the probability and consequences of an adverse outcome (29). Our analyses suggest that an increasing total of active COVID-19 cases is associated with higher levels of anxiety confirming the assumptions of Reardon et al who stated that the COVID-19 pandemic could cause anxiety in elite athletes, which was confirmed by our findings (30). Addressing anxiety in athletes is particularly relevant as their livelihood depends on their ability to perform in training and competition. Other factors than the COVID-19 pandemic are driving anxiety in elite athletes, for example career dissatisfaction, fear of musculoskeletal injury or increasing age (31). Therefore, those responsible for the psychological care of athletes should be aware of anxiety related to the COVID-19 pandemic as it adds to the existing anxiety burden. However, in the current study, the increasing burden of anxiety during the ongoing pandemic was even more pronounced in staff members than in Football players and their need for mental wellbeing should not be neglected, either.

In our study, we employed two distinct indicators of COVID-19 incidence (i.e. novel and active COVID-19 cases). Surprisingly, we found that associations of these two indicators with the different mental health proxies were not comparable. Novel cases of COVID-19 were only significantly associated with worse sleep quality, while active cases of COVID-19 were significantly associated with increasing levels of perceived infection risk and location variance. Potential explanations for this phenomenon might be found in the media coverage on both concepts during the observation period.

During the latter, the number of novel cases per day was small and remained largely unaltered (23). Therefore, it seems plausible that no alterations are found in the levels of anxiety and location variance. Media coverage on novel COVID-19 cases might have led to a higher level of cogitation on the implications of this information resulting in disturbances in sleep quality. The number of active cases also included cases that were diagnosed prior to the observation period and was naturally higher than the number of novel cases. Consequently, the confrontation with such numbers might have caused enhanced feelings of anxiety.

### Strengths and limitations

This study offered the unique possibility to study various mental health outcomes in a population of elite Footballers providing insight in their response to an increasing COVID-19 incidence. The combination of data from multiple sources (i.e. governmental data on COVID-19 incidence, GNSS-data from a specifically designed app and self-reported mental health data) in the statistical analyses of our paper emphasize the individuality and quality of the current study. However, several limitations should be addressed. The data collected by the smartphone app were fragmented and sampling unbalanced (i.e. largest proportion of observations was affiliated with club E) leading to potential misinterpretation of the association of COVID-19 incidence with location variance. The lack of data density may be attributable to participants reluctance to continuous submission of location data (14). We aimed to mitigate this issue by aggregating the data, relying on solid statistical procedures and cautiously drawn conclusions. The current study collected data during a rather stable period in the COVID-19 pandemic (23). It is likely, that more variance in the mental health proxies would have been observed 1) in a period with more rapidly growing COVID-19 incidence or 2) when employing instruments that are more sensitive to small changes in mental health. Yet, it should be noted that the original study (from which the data for the current analyses were derived) did not specifically address mental health changes but rather focused on the implementation of a sound monitoring concept for safe resumption of professional Football. On this line, we aimed for ideal processing of the data at hand. The current study yielded interesting findings that deserve further exploration and may aid in the monitoring and handling of mental health in professional sports during the ongoing and potential future pandemics.

## Conclusion

An increasing COVID-19 incidence is adversely associated with mental health in professional Footballers and staff members of their team. This emphasizes the need for additional, targeted mental care during the pandemic, which can be included in the daily routines of this population.

## DECLARATIONS/COMPLIANCE WITH ETHICAL STANDARDS

### Licence statement

I, the Submitting Author has the right to grant and does grant on behalf of all authors of the Work (as defined in the below author licence), an exclusive licence and/or a non-exclusive licence for contributions from authors who are: i) UK Crown employees; ii) where BMJ has agreed a CC-BY licence shall apply, and/or iii) in accordance with the terms applicable for US Federal Government officers or employees acting as part of their official duties; on a worldwide, perpetual, irrevocable, royalty-free basis to BMJ Publishing Group Ltd (“BMJ”) its licensees and where the relevant Journal is co-owned by BMJ to the co-owners of the Journal, to publish the Work in British Journal of Sports Medicine and any other BMJ products and to exploit all rights, as set out in our <u>licence</u>.

The Submitting Author accepts and understands that any supply made under these terms is made by BMJ to the Submitting Author unless you are acting as an employee on behalf of your employer or a postgraduate student of an affiliated institution which is paying any applicable article publishing charge (“APC”) for Open Access articles. Where the Submitting Author wishes to make the Work available on an Open Access basis (and intends to pay the relevant APC), the terms of reuse of such Open Access shall be governed by a Creative Commons licence – details of these licences and which <u>Creative Commons</u> licence will apply to this Work are set out in our licence referred to above

## Data Availability

Data included in the manuscript can be made available upon reasonable request.

## Authors contributions

All authors contributed to the study conception and design. Material preparation, data collection and analysis were performed by Alexander Seymer, Antje van der Zee-Neuen and Dagmar Schaffler-Schaden. The first draft of the manuscript was written by Antje van der Zee-Neuen and all authors commented on previous versions of the manuscript. All authors read and approved the final manuscript.

## Ethics/Informed consent

The current study presents secondary analyses of data collected in the context of the scientific surveillance of five selected, professional Austrian Football clubs.

The study protocol of the initial study and all procedures were approved by the Austrian ethics committee of Salzburg county (statement of the ethics board of Salzburg county, ID 415-EP/73/820-2020). Informed consent was obtained from all individual participants involved in the study. Participants were informed about the study purpose and procedures in writing and were additionally informed in person prior to providing their written consent.

## Guidelines and regulations

All procedures performed in the study were in accordance with the ethical standards of the Austrian ethics committee of Salzburg county and with the 1964 Helsinki Declaration and its later amendments.

## References

1. Galea S, Merchant RM, Lurie N. The Mental Health Consequences of COVID-19 and Physical Distancing: The Need for Prevention and Early Intervention. JAMA Intern Med. 2020;180(6):817–8.

2. Rajkumar RP. COVID-19 and mental health: A review of the existing literature. Asian J Psychiatr. 2020;52:102066.

3. Hawryluck L, Gold WL, Robinson S, Pogorski S, Galea S, Styra R. SARS control and psychological effects of quarantine, Toronto, Canada. Emerg Infect Dis. 2004;10(7):1206–12.

4. Hossain MM, Sultana A, Purohit N. Mental health outcomes of quarantine and isolation for infection prevention: a systematic umbrella review of the global evidence. Epidemiol Health. 2020;42:e2020038.

5. Brooks SK, Webster RK, Smith LE, Woodland L, Wessely S, Greenberg N, et al. The psychological impact of quarantine and how to reduce it: rapid review of the evidence. Lancet. 2020;395(10227):912–20.

6. Rehm J, Shield KD. Global Burden of Disease and the Impact of Mental and Addictive Disorders. Curr Psychiatry Rep. 2019;21(2):10.

7. Mumba MN, Nacarrow AF, Cody S, Key BA, Wang H, Robb M, et al. Intensity and type of physical activity predicts depression in older adults. Aging Ment Health. 2020:1–8.

8. Teychenne M, White RL, Richards J, Schuch FB, Rosenbaum S, Bennie JA. Do we need physical activity guidelines for mental health: What does the evidence tell us?: Mental Health and Physical Activity; 2020.

9. van der Zee-Neuen A, Wirth W, Hoesl K, Osterbrink J, Eckstein F. The association of physical activity and depression in patients with, or at risk of, osteoarthritis is captured equally well by patient reported outcomes (PROs) and accelerometer measurements - Analyses of data from the Osteoarthritis Initiative. Seminars in Arthritis and Rheumatism.2019;49(3): 325–30.

10. Gulliver A, Griffiths KM, Christensen H. Barriers and facilitators to mental health help-seeking for young elite athletes: a qualitative study. BMC Psychiatry. 2012;12:157.

11. Souter G, Lewis R, Serrant L. Men, Mental Health and Elite Sport: a Narrative Review. Sports Med Open. 2018;4(1):57.

12. Saeb S, Zhang M, Karr CJ, Schueller SM, Corden ME, Kording KP, et al. Mobile Phone Sensor Correlates of Depressive Symptom Severity in Daily-Life Behavior: An Exploratory Study. J Med Internet Res. 2015;17(7):e175.

13. Canzian L, Musolesi M. Trajectories of depression: unobtrusive monitoring of depressive states by means of smartphone mobility traces analysis. Proceedings of the 2015 ACM international joint conference on pervasive and ubiquitous computing. 2015. p. 1293–304

14. van der Zee-Neuen A, Schaffler-Schaden D, Herfert J, O Brien J, Johansson T, Kutschar P, et al. Team contact sports in times of the COVID-19 pandemic-a scientific concept for the Austrian football league. medRxiv; 2020.

15. João KADR, Jesus SN, Carmo C, Pinto P. The impact of sleep quality on the mental health of a non-clinical population. Sleep Med. 2018;46:69–73.

16. Kellmann M, Bertollo M, Bosquet L, Brink M, Coutts AJ, Duffield R, et al. Recovery and Performance in Sport: Consensus Statement. Int J Sports Physiol Perform. 2018;13(2):240-5.

17. Komoot P. https://photon.komoot.io [

18. BING. https://docs.microsoft.com/en-us/bingmaps/rest-services/ [

19. platform C-D. Austrian National Public Health Institute (Gesundheit Österreich GmbH, GÖG). 2020.

20. Souza RA, Beltran OAB, Zapata DM, Silva E, Freitas WZ, Junior RV, et al. Heart rate variability, salivary cortisol and competitive state anxiety responses during pre-competition and pre-training moments. Biol Sport. 2019;36(1):39–46.

21. Team RC. R: A language and environment for statistical computing. R Foundation for Statistical Computing, Vienna, Austria; 2020.

22. Bruns DP, Kraguljac NV, Bruns TR. COVID-19: Facts, Cultural Considerations, and Risk of Stigmatization. J Transcult Nurs. 2020;31(4):326–32.

23. AGES. Austrian Agency for Health and Food Safety, Epidemiological Parameters of the COVID-19 Pandemic (Epidemiologische Parameter des COVID19 Ausbruchs), Updates June-July. 2020.

24. Le H, Schmidt FL, Putka DJ. The multifaceted nature of measurement articfacts and its implications for estimating construct-level relationships. Organizational Research Methods; 2007. p. 165–200.

25. Gupta R, Grover S, Basu A, Krishnan V, Tripathi A, Subramanyam A, et al. Changes in sleep pattern and sleep quality during COVID-19 lockdown. Indian J Psychiatry. 2020;62(4):370–8.

26. Fu W, Wang C, Zou L, Guo Y, Lu Z, Yan S, et al. Psychological health, sleep quality, and coping styles to stress facing the COVID-19 in Wuhan, China. Transl Psychiatry. 2020;10(1):225.

27. Tayech A, Mejri MA, Makhlouf I, Mathlouthi A, Behm DG, Chaouachi A. Second Wave of COVID-19 Global Pandemic and Athletes’ Confinement: Recommendations to Better Manage and Optimize the Modified Lifestyle. Int J Environ Res Public Health. 2020;17(22).

28. Mandelkorn U, Genzer S, Choshen-Hillel S, Reiter J, Meira E Cruz M, Hochner H, et al. Escalation of sleep disturbances amid the COVID-19 pandemic: a cross-sectional international study. J Clin Sleep Med. 2020.

29. Sjöberg L. Factors in risk Perception. Risk Analysis; 2002. p. 1–12.

30. Reardon CL, Bindra A, Blauwet C, Budgett R, Campriani N, Currie A, et al. Mental health management of elite athletes during COVID-19: a narrative review and recommendations. Br J Sports Med. 2020.

31. Rice SM, Gwyther K, Santesteban-Echarri O, Baron D, Gorczynski P, Gouttebarge V, et al. Determinants of anxiety in elite athletes: a systematic review and meta-analysis. Br J Sports Med. 2019;53(11):722–30.

