## Supplementary for "Association of COVID-19 incidence with objectively and subjectively measured mental health proxies in the Austrian Football League – an epidemiological study"

| *Table S1: Overview of local administrative districts included in the current study* | |
| --- | --- |
| *1* | *Burgenland-North* |
| *2* | *Burgenland-South* |
| *3* | *Graz* |
| *4* | *Innviertel* |
| *5* | *Carinthia-East* |
| *6* | *Carinthia-West* |
| *7* | *Liezen* |
| *8* | *Mostviertel* |
| *10* | *Mühlviertel* |
| *11* | *Lower Austria-Center* |
| *12* | *Upper Austria Central Area Linz* |
| *13* | *Upper Austria Central Area Wels* |
| *14* | *Eastern Upper Styria* |
| *15* | *Eastern Styria* |
| *16* | *East Tyrol* |
| *17* | *Pinzgau/Pongau/Lungau* |
| *18* | *Pyhrn-Eisenwurzen* |
| *19* | *Rhine-Valley/Bregenz Forest* |
| *20* | *Salzburg-North* |
| *21* | *Thermenregion* |
| *22* | *Tyrol North-East* |
| *23* | *Tyrol-West* |
| *24* | *Tyrol- Central Area* |
| *25* | *Traunviertel/Salzkammergut* |
| *26* | *Vorarlberg-South* |
| *27* | *Waldviertel* |
| *28* | *Weinviertel* |
| *29* | *Western/Southern Styria* |
| *30* | *Western Upper-Styria* |
| *31* | *Vienna Central/Southeast* |
| *32* | *Vienna Northeast* |
| *33* | *Vienna-West* |

**Figure S1: Detailed illustration of distribution of GNSS data points per user**

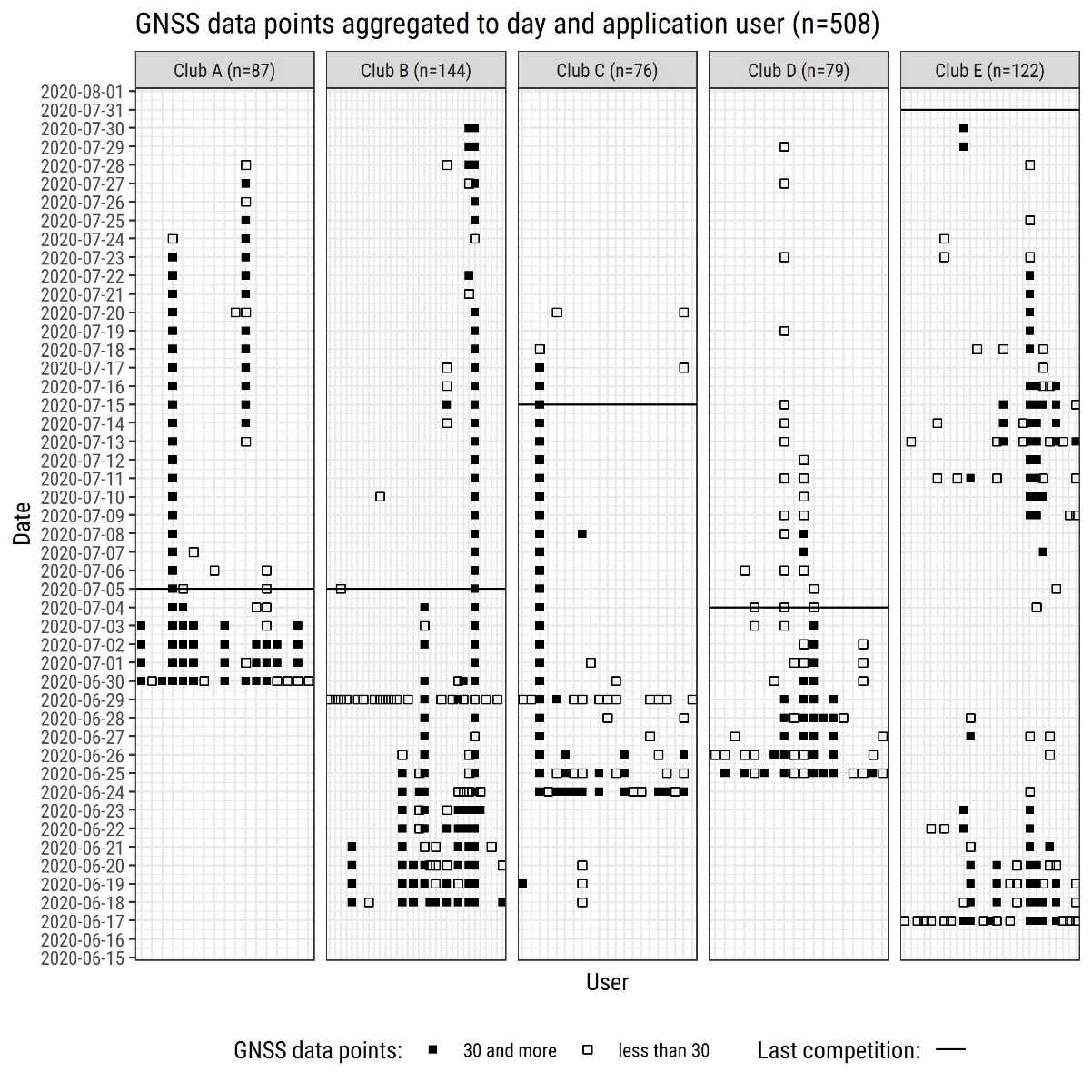

**Figure S2: Correlation matrix of numeric parameters including distribution plots (pairwise deletion).**

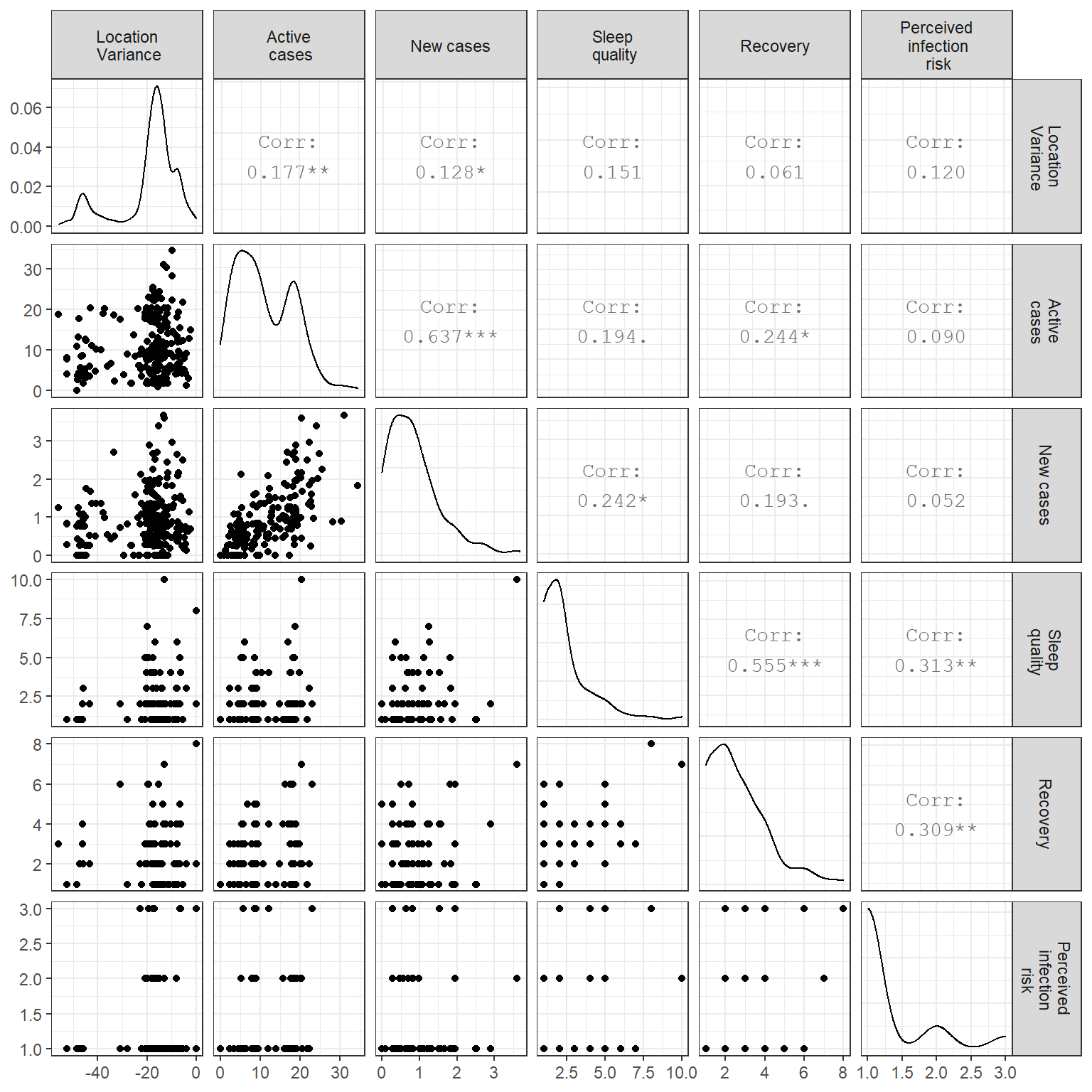
